# PhenoID, a language model normalizer of physical examinations from genetics clinical notes

**DOI:** 10.1101/2023.10.16.23296894

**Authors:** Davy Weissenbacher, Siddharth Rawal, Xinwei Zhao, Jessica R. C. Priestley, Katherine M. Szigety, Sarah F. Schmidt, Mary J. Higgins, Arjun Magge, Karen O’Connor, Graciela Gonzalez-Hernandez, Ian M. Campbell

## Abstract

**Background:** Phenotypes identified during dysmorphology physical examinations are critical to genetic diagnosis and nearly universally documented as free-text in the electronic health record (EHR). Variation in how phenotypes are recorded in free-text makes large-scale computational analysis extremely challenging. Existing natural language processing (NLP) approaches to address phenotype extraction are trained largely on the biomedical literature or on case vignettes rather than actual EHR data.

**Methods:** We implemented a tailored system at the Children’s Hospital of Philadelpia that allows clinicians to document dysmorphology physical exam findings. From the underlying data, we manually annotated a corpus of 3136 organ system observations using the Human Phenotype Ontology (HPO). We provide this corpus publicly. We trained a transformer based NLP system to identify HPO terms from exam observations. The pipeline includes an extractor, which identifies tokens in the sentence expected to contain an HPO term, and a normalizer, which uses those tokens together with the original observation to determine the specific term mentioned.

**Findings:** We find that our labeler and normalizer NLP pipeline, which we call PhenoID, achieves state-of-the-art performance for the dysmorphology physical exam phenotype extraction task. PhenoID’s performance on the test set was 0.717, compared to the nearest baseline system (PhenoTagger) performance of 0.633. An analysis of our system’s normalization errors shows possible imperfections in the HPO terminology itself but also reveals a lack of semantic understanding by our transformer models.

**Interpretation:** Transformers-based NLP models are a promising approach to genetic phenotype extraction and, with recent development of larger pre-trained causal language models, may improve semantic understanding in the future. We believe our results also have direct applicability to more general extraction of medical signs and symptoms.

**Funding:** US National Institutes of Health

## 1 Introduction

The practice of clinical genetics aims to determine a genetic etiology of an individual’s medical presentation. This process involves clinical correlation of the individual’s genetic variants with their “phenotype”–their physical, physiologic, and functional characteristics. A critical component of genetic phenotyping is the dysmorphology physical examination which specifically catalogues morphological differences of the patient’s facial structure or body. However, more general medical signs such as neurologic dysfunction may also be identified during the examination. The findings directly influence clinical diagnosis, the selection of genetic testing, and the interpretation of results. Phenotype correlation is particularly important when genetic tests reveal variants of uncertain clinical significance. Beyond the clinic, phenotypic information is also useful to researchers attempting to delineate undescribed genetic conditions and to further the understanding of existing conditions. Despite the specific terminology relevant to genetics, phenotyping is analogous to the collection of signs and symptoms in general medical practice.

Physical examinations, including dysmorphology exams, are frequently documented in the electronic health record (EHR) as a series of organ system observations (Figure 2.1, right panel). To standardize the description and comparison of phenotypic findings, many clinicians and laboratory professionals refer to the Human Phenotype Ontology (HPO) [1]. This ontology is specially designed for human genetics. The coding (annotation or labeling) of documented findings into HPO terms is highly labor-intensive and frequently requires advanced training in genetics for the best results. Because findings are captured in the EHR as unstructured free text, proper identification of the mentions and their mapping (normalization) to HPO terms is essential for downstream computational analysis. Thus, automating the annotation process is needed in order to conduct studies at scale and infer relevant clinically-relevant patterns.

**Figure 1:**
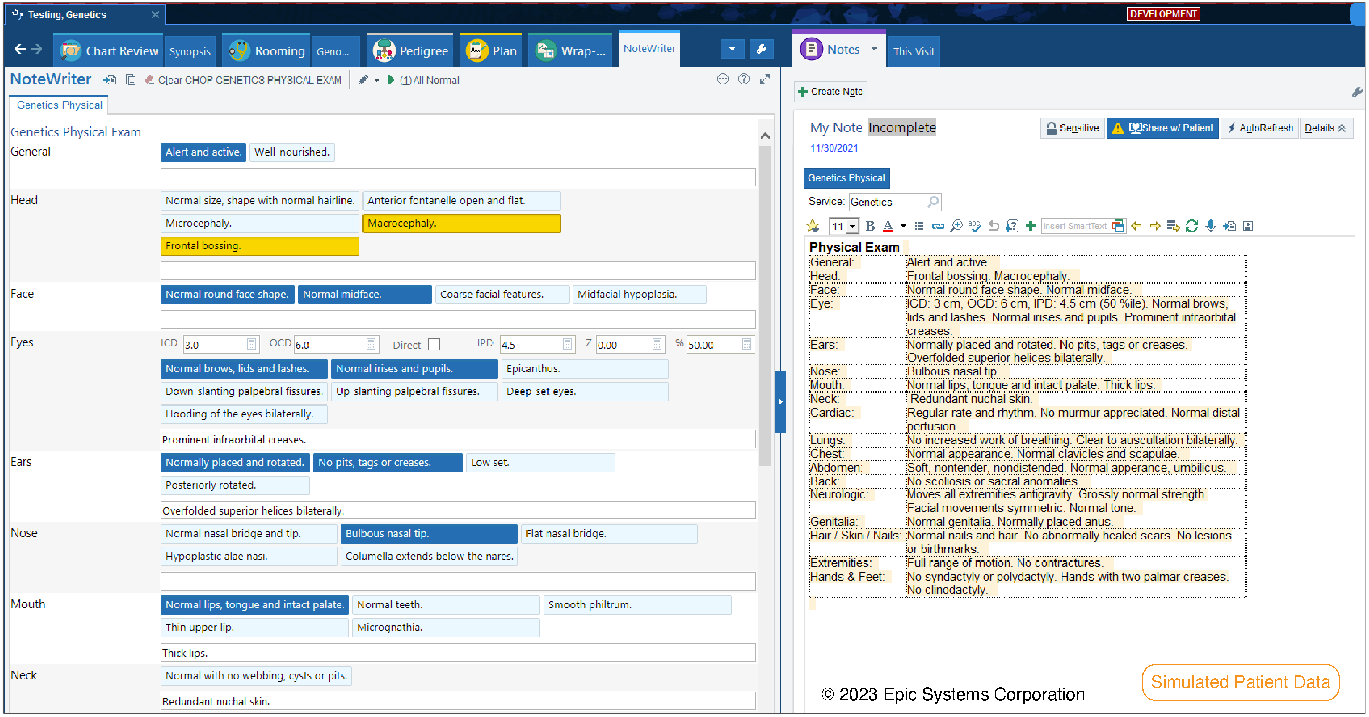
EHR Dysmorphology Interface with Simulated Data. Left: The system contains buttons for common dysmorphology physical exam finding organized by organ system with accompanying free text boxes for open-ended documentation. Functionality is provided for automatic calculation of specialized body measurements. Right: Clinical text is then automatically generated based on the combination of buttons pressed and free text entered. The buttons map to HPO terms.

**Figure 2:**
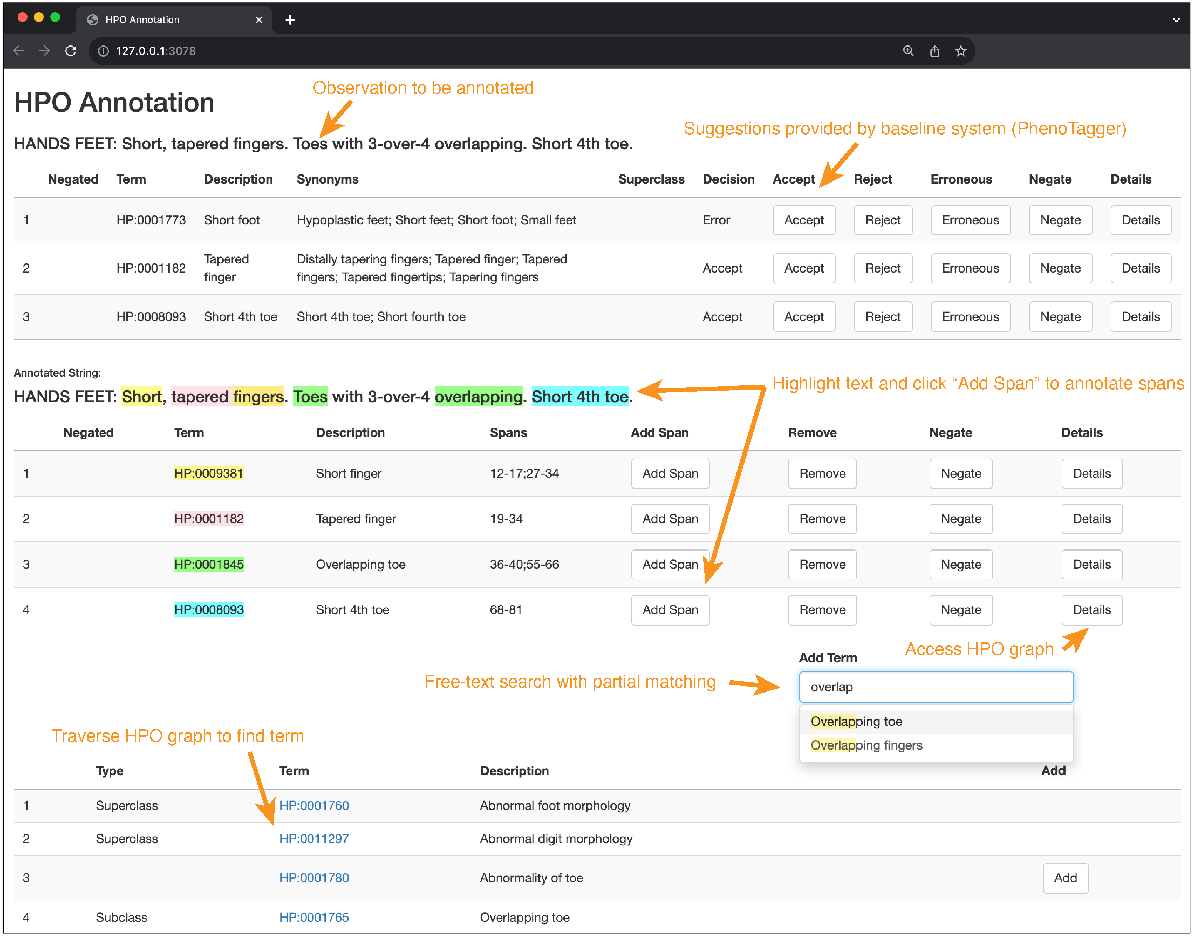
Custom Annotation Interface. Top: The clinical observation to be annotated is presented along with suggested annotations precomputed with one of the baseline systems, PhenoTagger. Functionality is provided to accept, revise, negate (PhenoTagger provides no negation detection) or reject each suggestion. Center: The same observation text is provided again to facilitate span annotation. A free-text search box with partial string matching allows selection of additional terms. Once the appropriate terms are selected, portions of the text are highlighted and can then be annotated to a particular term. Bottom: Functionality is provided to traverse the HPO graph to find a more appropriate hyperonym or hyponym.

Advanced natural language processing (NLP) methods can reduce the cost of retrieving and standardizing mentions of phenotypes from the EHR. In this study, we propose a pipeline to extract and normalize the key phenotypic findings documented in dysmorphology physical examinations. The extraction step is generally challenging due to the descriptive style of the examinations and their polarity (positive or negative findings). Further complications arise given that the observations are short reports where, for concision, practitioners often mention two findings with disjoint and overlapping phrases. Standard sequence labeling approaches are designed to extract contiguous and mutually exclusive terms and often fail to capture disjoint and overlapping terms[2]. As an additional challenge, our labeler should also resolve the polarity of the findings to return only the abnormal findings and ignore normal ones.

Consider for example the following organ system observation: “`EYES: long palpebral fissures with slight downslant. Normal eyebrows.`” Two dysmorphology phenotypes are discussed: *a. Long palpebral fissure - HP:0000637* and *b. Downslanted palpebral fissures - HP:0000494*. Our labeler should *extract* the span of the text referring to the phenotypes *long palpebral fissures* and *palpebral fissures with slight downslant* and then *normalize* them to the term IDs of the corresponding terms in the HPO, `HP:0000637` and `HP:0000494`, respectively. However, the text *palpebral fissures* contributes to both the `HP:0000637` and `HP:0000494` terms, and is thus an *overlapping mention*, which our labeler handles. Our labeler will ignore normal findings (such as “Normal eyebrows”). Consider also an example of a disjoint finding (whereby the finding is defined with non-consecutive words): `NOSE: Short, wide nasal bridge. Anteverted nares.`” This should be labeled as `Short nasal bridge - HP:0003194`

The normalization step is challenging due to both the large scale of the HPO ontology and the inconsistent levels of term detail available. With more than 17,000 terms and with most terms having synonyms, the HPO ontology is a very large knowledge base. An automatic normalizer would necessarily learn with very incomplete data, as any manually annotated training corpus will be limited in size and will provide examples of only a small percentage of the terms in the HPO that occur in the context of real observations. Furthermore, while specifically designed for human genetics and constantly improving, the HPO does not have standardized levels of term detail. Therefore, a phenotypic finding may need to be matched with a close ancestor in the hierarchy of the ontology, making a typical string matching strategy for normalization very inefficient since the string of the ancestor will differ from the string of the key finding. For example, there exist both `Naevus flammeus of the eyelid - HP:0010733` and `Nevus flammeus of the forehead - HP:0007413`, but no such term for the nose, leaving only the generic `Nevus flammeus - HP:0001052` to normalize this abnormality of the nose.

The main contributions of our study are (1) a manually annotated corpus of observations from dysmorphology physical examinations with normal and dysmorphic findings; (2) a detailed evaluation of a pipeline relying on transformers to detect and normalize the dysmorphic findings achieving state-of-the-art performance; (3) and a comparison with competing phenotype normalizers.

## 2 Materials and methods

### 2.1 Data collection and annotation

In order to better capture the physical manifestations of genetic disease in our practice, we built a standardized system to document the findings in our EHR (Epic Systems Inc.) [3]. The specialized data entry form, shown in Figure 2.1, uses the vendor’s standard functionality (a SmartBlock SmartForm™) and can be inserted into any clinical note. It features buttons and text boxes linked to specific organ systems. Our primary goal when building this system was to facilitate the structured extraction of physical examination findings. We mapped individual button responses to HPO terms; for example the “Thin upper lip” button in the “Mouth” system corresponds to HP:0000219. The extraction of findings from the free text responses was a primary impetus for the NLP system.

The Institutional Review Board at Children’s Hospital of Philadelphia determined that this study met exemption criteria under 45 CFR 46.104(d)4(iii) as protocol 22-019752. A waiver of HIPAA authorization under 45 CFR 164.512(i)(2)(ii) was granted to access identifiable information from the medical records.

In April 2022, we accessed the underlying data structure of the physical exam system (the SmartData Elements™) and retrieved all free text responses. There were 34 distinct authors, with 24 authors having written at least 10 observations. Many textual responses were identical between individuals. This is likely primarily a function of users creating macros which include the same text automatically; however, some observations may have been manually repeated. The number of repeats are included in the data, but no extra weight was included during model training. Then, we de-identified the observations using NLM Scrubber [4] to remove potentially identifiable health information annotated each observation. To speed up annotation, we pre-computed potential annotations with a baseline system, PhenoTagger [5]. We developed a custom annotation interface, shown in Figure 2.1, to incorporate in the interface the pre-annotations of PhenoTagger and quickly traverse the ontology graph to identify the most appropriate term when correcting the pre-annotations or labeling missing terms.

We instructed our annotators to identify all phenotypes unambiguously documented in the text from the 2022-06-11 release of HPO. This includes abnormal findings as well as specifically mentioned normal findings. Because the HPO does not include terms for normal findings, the closest matching abnormal finding was used and indicated as negated.

After the HPO term annotation, the annotators would select the spans of text that unambiguously determine the HPO term. There were 4 annotators, all physicians in various stages of Clinical Genetics training at the time of annotation. One was an attending physician and three were Pediatrics / Genetics residents. We annotated 890 observations at least twice. Overall, there was complete inter-annotator agreement of all positive HPO terms for 76.1% of observations. Among observations with zero, one or two positive HPO terms mentioned, there was complete concordance for 85.5% of these observations. However, for observations with three to eight positive HPO terms mentioned, there was only 47.5% concordance, underscoring the challenging nature of the annotation task. In addition to additive error chance per term, we hypothesize that longer observations are distracting. Average F1 score for annotators when compared to all permutations of other annotators was 0.844. Discrepancies were automatically adjudicated by selecting the ones of the annotator with the most years of clinical experience.

### 2.2 Corpus

Our annotation process resulted in 3826 (3136 unique) organ system - observation textual entries documented for 1655 unique individuals. A total of 935 HPO terms were annotated at least once. The EHR Interface (2.1) contained buttons for 22 commonly observed HPO terms. This made those terms much less likely to be described in the free text. Despite this, 362 observations contained at least one of these 22 terms, either because the finding required additional details or the clinician was unaware of the button.

### 2.3 PhenoID, a proposed normalization pipeline

We aim to learn a function that takes an observation and assigns all HPO term IDs to the dysmorphic findings mentioned in the observation. The function should allow the repetitions of the same HPO term ID in the list if the HPO term is mentioned multiple times in the observation. It should return an empty list if no dysmorphic finding was mentioned in the observation.

Our first choice of architecture to design the normalizer was a multi-label classifier, which is a natural choice to resolve this task. However, we found difficult to train a multi-label classifier on our data, achieving very poor performance during our preliminary experiments. As an alternative, we propose a pipeline to learn the function. Figure 3 summarizes its architecture, which we detail next.

**Figure 3:**
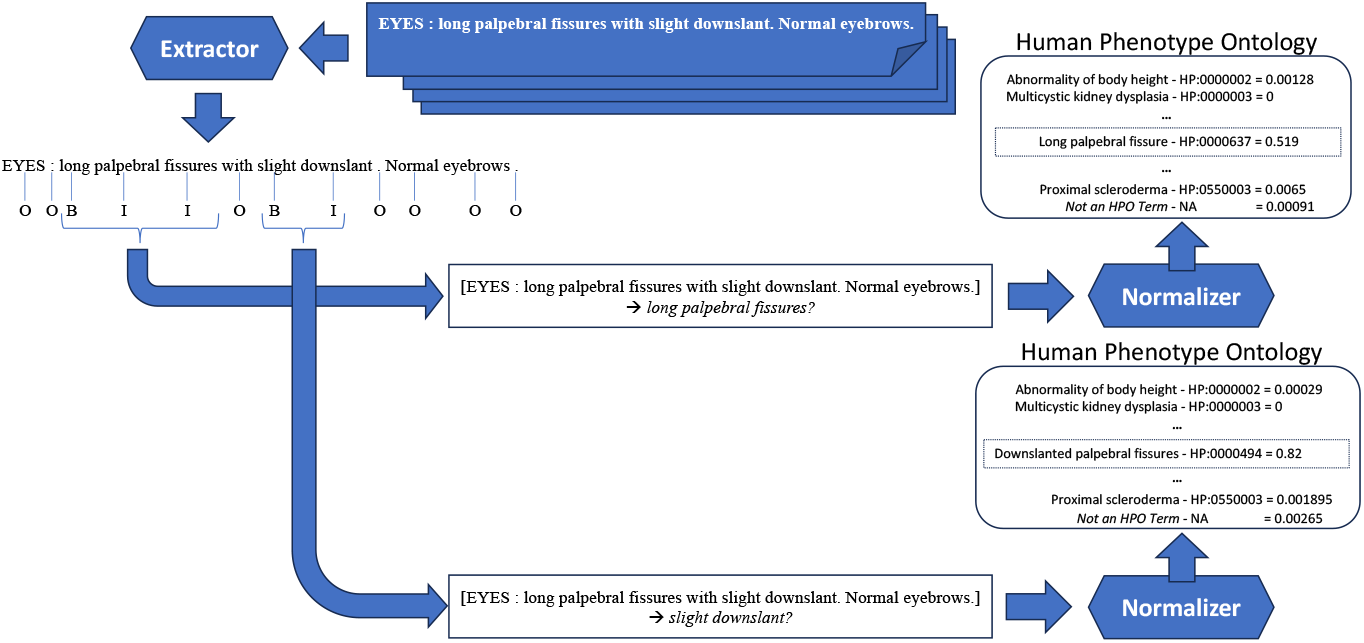
PhenoID. Given an observation, the extractor predicts the probabilities for each token of the observation to **B**egin, to be **I**nside, or **O**utside a mention of a dysmorphic finding. Each mention detected by the extractor is then passed, one at a time and along with the text of the observation, to the normalizer which predicts the most probable HPO term ID denoted by the mention, or the special value *NA* if the mention extracted is more likely not an HPO term or is a normal finding.

Our pipeline, *PhenoID*, is composed of two modules. The extractor module is a sequence labeler. It focuses only on detecting the spans of the mentions of dysmorphic findings in an observation while ignoring the mentions of normal findings. The normalizer module takes as input the text of the observation and the spans of *one* dysmorphic finding detected by the extractor. It is responsible for mapping the given dysmorphic finding to the most specific HPO term ID referencing the finding. Each module relies on transformers to perform their tasks[6]. Our pipeline has two main advantages over a multi-label classifier. The pipeline architecture allows us to decouple the detection of the dysmorphic findings from their normalization. This helps both neural networks to learn the best set of features for each task and optimize independently and efficiently their loss functions. Once extracted, since the candidate terms are presented one at a time to the normalizer, the normalizer can focus its attention mechanism to learn features encoding the semantic properties that are specific to the only candidate term presented and to the context where it appears.

#### 2.3.1 Extraction

We opted for the currently dominant architecture for NLP systems, a transformer architecture [7]. It encodes the meaning of a sentence into a multidimensional space to learn specific syntactic or semantic relations between the words composing the input text. These relations are used as features by the last layer of the classifier, a feed-forward layer, to predict the label of each token of the observation to be inside or outside a mention of an HPO term. With the increasing popularity of the transformer models, the community released multiple transformers pre-trained with different algorithms and/or on different corpora. For our experiments, we chose the Bio-ClinicalBERT pre-trained transformer from the Hugging Face repository. This model was initialized with a general BERT pre-trained model which was further trained on PubMed abstracts and PubMed central full articles, and then on all de-identified clinical notes from the MIMIC-III database [8]. We opted for this model to ensure it has a representation for the most common medical concepts and a basic knowledge of their relationships.

We formulated the detection of HPO term mentions as a sequence labeling problem [2]. In this approach, given an observation, our transformer estimates the probabilities for each token to begin a phrase mentioning an HPO term by predicting the label *B*, or to be inside the phrase by predicting the label *I*, or to be outside any HPO term mentions by predicting the label *O*. This is known as the BIO schema.

#### 2.3.2 Normalization

We also opted for the transformer architecture to implement our normalizer and fine-tuned a Bio-ClinicalBERT pretrained model. We formulated the normalization task has a classification task. We gave as input to the transformer the span of one HPO term to normalize and the text of the observation where the span was found. The transformer predicts a probability distribution over 11,988 HPO term IDs. We normalized the HPO term with the HPO term ID for which the transformer assigned the highest mass probability. The Human Phenotype Ontology has 16,908 term IDs at the time of writing but not all terms listed in the ontology are observable during a dysmorphology physical examination, for example *HP:0000089, Renal hypoplasia* or *HP:5000043, Anti-D2 R antibody*. We removed those term IDs from the IDs considered by our normalizer. We ensured that our training set contained at least one example of each observable HPO term. To do so, we generated new examples simply composed of a preferred term or a synonym. For example, for the HPO term ID *HP:0000185*, we generated four new examples, the example *Cleft soft palate* for the preferred term and the example *Cleft velum* for one of its synonym. We also over-sampled half of the new generated examples, by randomly selecting the terms to duplicate. Our final set of training examples has a total of 116,644 examples, with 113,877 generated examples added to our initial set of 2,767 annotated training examples (corresponding to 1,716 unique observations).

Note that the BIO schema was designed for labeling terms that are continuous and exclusive to each other. Thus, we could not represent disjoint or overlapping HPO terms with the BIO schema. Instead, we used two heuristics which are contrasted in Figure 4. In the first heuristic, the *noisy-BIO schema*, we applied the BIO schema to our training data, knowing that we will create incorrect and incomplete annotations for all disjoint/overlapping HPO terms. In the second heuristic, the *recast-BIO schema*, we applied the BIO schema to our training data but applied rules to correct the resulting annotations, accepting the loss of information after these corrections.

**Figure 4:**
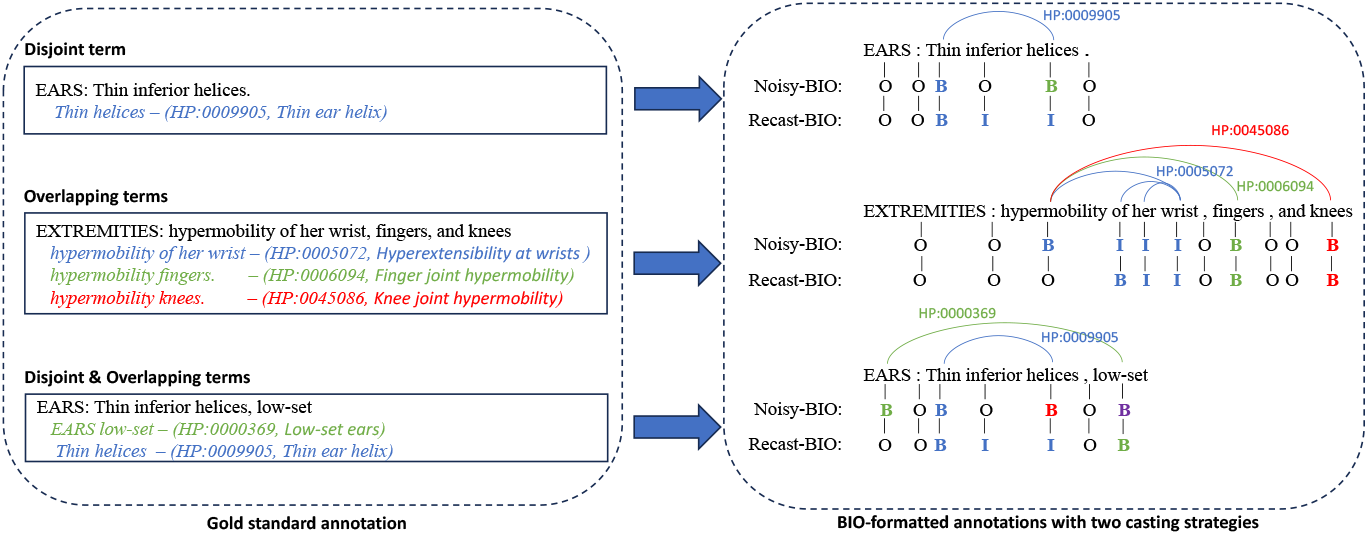
Noisy-BIO and Recast-BIO schemas applied on disjoint and overlapping HPO terms

The rationale behind our Noisy-BIO and Recast-BIO heuristics is that the detection of the dysmorphic findings is just an intermediary step needed to enable computation from the normalizer. The detection of the spans of the HPO term is not inherently a critical clinical research problem. We hypothesized that the normalizer would learn to ignore dependent tokens when presented without their heads by predicting *NA* and when only presented with the head of an HPO term learn to normalize it correctly by retrieving the dependent tokens from the text of the observation given as additional input. For instance, in Figure 4, the HPO term *Thin helices* is mentioned with the disjoint spans *Thin* - *helices*. We created two term mentions, *Thin* and *helices*, when we applied the Noisy-BIO schema to the annotation of the term. The normalizer should predict the special value *NA* when presented with the dependent *Thin* and predict the HPO ID *HP:0009905* when presented with the head *helices* and the text of the observation, retrieving from the text the dependent token *Thin* needed to do the prediction.

#### 2.3.3 Baselines

We evaluated PhenoID by comparing its performance with the performance of six existing normalizers. We chose these normalizers because they implemented intuitive or current state-of-the-art approaches and their source code was available and functional. This section summarizes the competing systems.

##### txt2hpo

The approach of txt2hpo[9] is intuitive, often effective, and should be the first approach attempted for phenotype identification^•^. The system normalizes the HPO terms by matching word stems of an observation with the word stems of the HPO terms. If two or more HPO terms are matched for the same stems, a similarity distance is computed between embeddings representing the tokens of the HPO terms and embeddings of the tokens of the observation. Then, the most similar HPO term is selected. The authors chose RegexpStemmer[10] from the NLTK library for their preprocessing and Doc2vec[11] from the gensim library[12] for computing the embeddings of the terms and the observations. The team provides optional functionality for the correction of the spelling within of the observation with a spellchecker and filtering of negated HPO terms using negSpacy[13].

##### Doc2HPO

Doc2HPO[14] performs a more complex string matching approach. It provides a customizable, human in the loop web interface to extract phenotype concepts from user provided texts and map these to HPO terms. The system allows the user to select from four parsing engines, including a simple string matching method and two previously developed systems, MetaMap and NCBO Annotator. Additionally, an ensemble of these parsing engines may be used that take the union of the results. Users of the system can be presented with the automated results where they can add, edit or accept them. Doc2HPO also incorporates negation detection using regular expressions. We compared our system with the string matching parse engine of Doc2HPO alone as well as with the ensemble of the four parsing engines.

##### Neural Concept Recoginition

Advancing beyond text matching, dictionary or ontology based look-up systems or other manual rule-based engineering methods, such as those used by NCBO Annotator or MetaMap, which achieve high precision but often low recall, Arbabi et al.[15] implemented a neural-based dictionary model, the Neural Concept Recognizer (NeuralCR), to predict if a word or phrase within a given text is synonymous with a concept in a reference ontology. The model encodes input text to vector representations using a convoluted neural network (CNN). The similarity of these representations are compared to embeddings learned from a representative ontology to identify relevant phrases for extraction. The embeddings from the ontology are learned by considering unique features compared to ancestor concepts, and the taxonomy’s structure is used to derive these representations. The model has some limitations such as not considering overlapping concepts.

##### PhenoTagger

Luo et al.[5] addresses the limitations of Neural Concept Recognizer with PhenoTagger, a hybrid method combining dictionary and machine learning methods. For their approach, a dictionary was constructed from the HPO which was used for the dictionary matching method and to build a training dataset using distant supervision. A BioBERT model was trained with this dataset to classify each candidate N-gram to an HPO concept, keeping concepts above the pre-determined threshold. The results of the dictionary-matching and the deep learning modules are then combined for the final output.

##### PhenoBert

While BERT models have improved SOTA performance on many NLP tasks, the computational time for classification is longer than CNN models. Feng et al.[16], proposed using a two level hierarchical CNN (TLH-CNN) models prior to implementing a BERT model. In their PhenoBERT pipeline, clinically relevant text segments (CTS) are extracted using a deep learning module and those not matched to an HPO term using dictionary-based matching are processed by the TLH-CNN module which computed candidate HPO terms for the CTS. These candidates are input in the BERT module, a pretrained BioBERT, for evaluation and final classification.

#### 2.3.4 Evaluation

We compared the abilities of the competing systems to correctly normalize the mentions of dysmorphic findings in the observations, regardless of their abilities to extract those mentions. We define a true positive as a mention of an HPO term labeled and normalized by an annotator as a dysmorphic finding in an observation that was correctly detected and normalized by a system. We defined a false positive as an HPO term that was incorrectly detected or normalized by a system - i.e. this HPO term was either not labeled by an annotator as being mentioned in the observation or the term was mentioned but it was negated. We defined a false negative as an HPO term mention in an observation that was labeled and normalized by an annotator but missed or normalized with a different HPO term ID by a system. Finally, we defined a true negative as the situation when both the annotator and normalizer indicated no HPO terms. We measured the performance on normalization with the standard F1-score, that is the harmonic mean of precision and recall. We also evaluated the performance of the systems when performing both extraction and normalization. In the exact extraction and normalization, we rewarded a system if it correctly detected at least part of the span of an affirmed HPO term mentioned in an observation and assigned the same HPO term ID as the annotator.

## 3 Results

We report the performance of all systems in Table 1. PhenoID achieved .717 F1 score on the normalization task, a score better than all scores of competing systems, showing the promise of our approach. In Table 2, we compare the performance of PhenoID with the two proposed BIO schemas. Whereas PhenoID outperforms all competing systems with both BIO schemas, interestingly, PhenoID with the Noisy-BIO schema achieved overall better performance than with the Recast-BIO schema. This result is rather counter intuitive. In the Recast-BIO heuristic, we trained PhenoID with truncated terms since we deleted outer tokens from the training examples of overlapping - and disjoint - terms, which makes the extractor more likely to detect the spans of these HPO terms partially. Despite the loss of information, this system achieved better recall than PhenoID with the Recast-BIO heuristic. On the contrary, in the Noisy-BIO heuristic, we trained PhenoID on spurious examples which were spans resulting from the split of labeled overlapping - and disjoint - terms into multiple spans. Regardless of these additional candidates detected by the extractor that the normalizer as to ignore or to normalize, PhenoID with the Noisy-BIO schema achieved better precision. Additional experiments with ad-hoc frameworks, such as the SHapley Additive exPlanations[17], are needed to explain the decisions taken by our models.

**Table 1:** Performance achieved by 7 competing systems when detecting and normalizing abnormal findings noted during dysmorphology physical examinations with their HPO Terms IDs

| System | Normalization only |  |  | Overlapping Extraction & Normalization |  |  |
| --- | --- | --- | --- | --- | --- | --- |
| | $F_1$ | P | R | $F_1$ | P | R |
| **PhenoID** | **.717** | **.736** | **.699** | **.715** | **.735** | **.696** |
| doc2HPO - String Matching | .515 | .6120 | .445 | .515 | .612 | .445 |
| doc2HPO - Ensemble | .573 | .727 | .472 | .573 | .727 | .472 |
| NeuralCR | .578 | .574 | .582 | .577 | .573 | .580 |
| PhenoBERT | .580 | .668 | .513 | .578 | .667 | .510 |
| PhenoTagger | .633 | .587 | .687 | .632 | .586 | .685 |
| txt2HPO | .616 | .700 | .549 | .615 | .700 | .548 |

**Table 2:** Comparison of PhenoID performance with different settings

| System | Normalization only |  |  | Overlapping Extraction & Normalization |  |  |
| --- | --- | --- | --- | --- | --- | --- |
| | $F_1$ | P | R | $F_1$ | P | R |
| PhenoID - Noisy-BIO | **.717** | **.736** | **.699** | **.715** | **.735** | **.696** |
| PhenoID - recast-BIO | .685 | .641 | **.735** | .684 | .641 | **.733** |
| *PhenoID (with gold truth spans given)* | .820 | .820 | .820 | - | - | - |

We also reported in Table2 the performance of PhenoID when the gold truth spans were given to the normalizer, which represents the hypothetical top performance of the normalizer if the extraction step output were perfect. We summarized the error categories along with examples in Table 3. We revealed the gold truth spans to the normalizer module of PhenoID to limit our analysis to the normalization errors without having to consider errors caused by a wrong extraction of the term spans. We randomly selected 105 observations where PhenoID incorrectly normalized an HPO term. After manual analysis, we identified ten categories of non-exclusive errors. Four categories account for the majority of errors (68.6%, 72). The most common errors a. (n = 26) are caused by imperfections of the HPO ontology itself. Such imperfections were terms that could reasonably be expected but are not defined (m = 17), synonyms terms listed with different IDs that should likely be merged (m = 11), or terms incorrectly defined (m = 1). Another common cause of errors b. is the presence of descriptive observation; clinicians describe the findings rather than providing the medical term present in the HPO. These descriptions often do not contain any words in common with the preferred terms. Despite the generalization allowed by the embeddings of the tokens, our neural network still seems to rely on keyword matching to retrieved these HPO terms. Also common, and often co-occurring with category b., are errors of category c. (n = 23) caused by a shift of the attention of the neural network to phrases outside the gold truth spans of the HPO term. Alternatively, the system seems to place too much attention given on an unrelated word. As an example, the neural network appeared to focused its attention on *profoundly* and not *dysmorphic* in the gold span and normalize *Child is profoundly dysmorphic* with *Profound global developmental delay - HP:0012736*. The errors of the last common category d. may not be considered as errors for some use cases since the neural network retrieved the hyperonyms of the HPO terms annotated. These terms are more general than the terms annotated by clinicians, however they are still accurate and closely related to the terms expected. The remaining categories of errors are marginal with less than 10 cases.

**Table 3:** Categories of normalization errors when gold truth spans are given to PhenoID

| Error type | count | Example |
| --- | --- | --- |
| a. Imperfect ontology | 24.8% (26) | NAILS HAIR SKIN: *hyperkeratosis on palms and soles* (diffuse not striate) [Palmoplantar hyperkeratosis - HP:0000972 labeled, Palmoplantar keratoderma - HP:0000982 synonym predicted] |
| b. Descriptive gold truth span | 19.0% (20) | EXTREMITIES: *Wind swept hands* [Ulnar deviation of finger - HP:0009465 labeled] |
| c. Misplaced attention | 12.4% (13) | GENERAL APPEARANCE: *Child is profoundly dysmorphic* has high flow nasal cannula in place. [Abnormal facial shape - HP:0001999 labeled, Profound global developmental delay - HP:0012736 predicted] |
| d. Hyperonym predicted | 12.4% (13) | GENITALIA: *Left inguinal cryptorchidism*. [Unilateral cryptorchidism - HP:0012741 labeled, Cryptorchidism - HP:0000028 predicted] |
| e. Misspelling | 5.7% (6) | EYES: *Long eyeashes*. Exotropia, normally wearing glasses. [Long eyelashes - HP:0000527 labeled] |
| f. Acronym | 4.8% (5) | NAILS HAIR SKIN: *jaundice appearance, hypertrichosis, sacral CDM* [Blue nevus - HP:0100814 labeled] |
| g. Hyponym predicted | 4.8% (5) | EYES: *Proptosis with conjunctival hemorrhage* of the right eye [Hemorrhage of the eye - HP:0011885 labeled, Subconjunctival hemorrhage - HP:0011896 predicted] |
| h. Complex mention requiring inference | 4.8% (5) | MOUTH: *Well-healed scar from the left upper lip to the nose from cleft lip/palate repair*. Gray retainer in place. Normal-appearing teeth. [Cleft lip - HP:0410030 labeled, Scarring - HP:0100699 predicted] |
| i. Unknown reason | 4.8% (5) | GENITALIA: *enlarged scrotum*; no hernia palpated [Abnormality of the scrotum - HP:0000045 labeled, Impacted tooth - HP:0011079 predicted] |
| j. Annotation error | 6.7% (7) | — |
| **TOTAL** | **100% (105)** |  |

We trained PhenoID by providing few examples of labeled observations and, for all preferred terms and their synonyms in the ontology, by generating examples containing just the terms themselves without contexts of use. The examples we generated are useful since they provide examples of the words composing the terms but, they provide no example of how one can refer to these terms by describing or explaining them. They also do not contain any information of the phrases likely to appear in their context and, more importantly, to phrases related or composing other HPO terms, causing the neural network to shift its attention to the words of the other terms during their resolution. We confirmed the lack of semantic understanding of the term descriptions and their contexts by looking at PhenoID’s predictions on a subset of terms in our test set. This subset is the set of terms occurring in the test set with no occurrences in the training set. This set is a well known challenge for normalizers. We analyzed the impact of the disjoint and overlapping terms for the predictions of our pipeline. Despite our simple representation of these terms, we found our pipeline able to detect and normalize many of them. We counted 146 disjoint terms in our test set and found that 67 (45.9%) were partially detected yet correctly normalized by our pipeline. Similarly, we counted 137 overlapping terms in our test set with 61 (44.5%) correctly normalized. The 70 terms in our test set that were both disjoint and overlapping were more difficult to normalize, with only 21 (30%) correctly normalized. We also analyzed the impact of negated terms and were surprised to find that among the 189 negated term (normal findings), only 3 (1.6%) terms were incorrectly predicted as abnormal findings by the extractor and presented to the normalizer. The extractor ignored, as expected, all other normal findings resolving almost perfectly this subtask.

## 4 Discussion

An important finding from our experiments was that, when we provided the gold truth spans to the normalizer it achieved a .82 F1 score, a score approaching human performance that we measured with the IAA .844 F1 score. This signals the need to improve the span extraction which currently limits the overall performance of our system with a lost of 10.3 points (0.82 - 0.717) as shown in Table 2 when comparing the performances achieved by PhenoID - Noisy-BIO and by PhenoID (with gold truth spans given). We are exploring the benefit of large language models (LLMs) to replace our sequence labeler with a generative model, whereby we present an observation in a prompt and ask the model to list all HPO terms mentioned in the observation as suggested in previous studies on different tasks[18].

Improving the span extraction is a promising direction, but the most important gain may come from improving the normalizer. When providing PhenoID with the gold truth spans of the terms, we achieved 0.82 F1 score, losing 18 points due to errors in normalization alone. We were unsuccessful during our first zero/few-shots experiments with chatGPT [19] for this purpose. ChatGPT was able to detect most of the HPO terms in the observations given, but it was not able to normalize them. It generated wrong or nonexistent HPO IDs. This signals the lack of understanding of the ontology and the need to fine-tune the model on medical knowledge bases to perform our specific task.

A possible limitation of our current dataset is the relative lack of diversity in the authors of our annotated data. While the observations were written by approximately 1.5% of all certified Clinical Geneticists in the United States [20], the algorithm may not perform well for text written by adjacent specialties (such as Clinical Biochemical Genetics) or at other institutions.

## 5 Conclusion

We implemented a purpose-built dysmorphology exam system in our EHR which allowed us to easily extract the text of physical exam observations. From the data generated by this system, we annotated a corpus of organ system observations with HPO terms using a custom web-based interface. We developed a two-step extractor / normalizer approach for the complex named entity recognition task mapping to the HPO. Our system achieves state-of-the-art performance on this task. An analysis of the errors of our system demonstrated some imperfections in the HPO ontology itself, but it also revealed a relative lack of semantic understanding by our model. We believe ongoing advances in NLP are likely to address this shortcoming and help generalize our system to other author specialties and documentation practices.

## Data Availability

All data produced in the present study are available upon reasonable request to the authors

https://github.com/Ian-Campbell-Lab/Clinical-Genetics-Training-Data/

## Author contributions

DW conceptualized the study, developed methodology, developed / programmed software, performed formal analysis, performed data curation, produced visualization, provided supervision and wrote the original draft of the manuscript. SR developed methodology, developed / programmed software, performed formal analysis and reviewed / edited the manuscript. XZ developed methodology, developed / programmed software, performed formal analysis, performed data curation and wrote the original draft of the manuscript. JRCP, KMS, SFS, and MJH performed formal analysis, performed data curation and reviewed / edited the manuscript. KO developed methodology, performed formal analysis, performed data curation and wrote the original draft of the manuscript. AM conceptualized the study, developed methodology, developed / programmed software, performed formal analysis, performed data curation and reviewed / edited the manuscript. GGH conceptualized the study, developed methodology, provided supervision, acquired funding and reviewed / edited the manuscript. IMC conceptualized the study, developed methodology, developed / programmed software, performed formal analysis, provided resources, performed data curation, produced visualization, provided supervision, acquired funding and wrote the original draft of the manuscript.

## Data availability statement

Newly annotated training data is available at https://github.com/Ian-Campbell-Lab/Clinical-Genetics-Training-Data/. Source code for the extractor and normalizer models are available at https://bitbucket.org/pennhlp/phenorm/.

## Footnotes

• At the time of writing, we were unable to locate an academic article describing txt2hpo, only the library was available to download on Github. We reverse-engineered its source code to understand the main approach of the authors.

## References

[1] Sebastian Köhler, Michael Gargano, Nicolas Matentzoglu, Leigh C. Carmody, David Lewis-Smith, Nicole A. Vasilevsky, Daniel Danis, Ganna Balagura, Gareth Baynam, Amy M. Brower, Tiffany J. Callahan, Christopher G. Chute, Johanna L. Est, Peter D. Galer, Shiva Ganesan, Matthias Griese, Matthias Haimel, Julia Pazmandi, Marc Hanauer, Nomi L. Harris, Michael J. Hartnett, Maximilian Hastreiter, Fabian Hauck, Yongqun He, Tim Jeske, Hugh Kearney, Gerhard Kindle, Christoph Klein, Katrin Knoflach, Roland Krause, David Lagorce, Julie A. Mc-Murry, Jillian A. Miller, Monica C. Munoz-Torres, Rebecca L. Peters, Christina K. Rapp, Ana M. Rath, Shahmir A. Rind, Avi Z. Rosenberg, Michael M. Segal, Markus G. Seidel, Damian Smedley, Tomer Talmy, Yarlalu Thomas, Samuel A. Wiafe, Julie Xian, Zafer Yüksel, Ingo Helbig, Christopher J. Mungall, Melissa A. Haendel, and Peter N. Robinson. The Human Phenotype Ontology in 2021. Nucleic Acids Research, 49(D1):D1207–D1217, January 2021.

[2] Fei Li, ZhiChao Lin, Meishan Zhang, and Donghong Ji. A span-based model for joint overlapped and discontinuous named entity recognition. In Proceedings of the 59th Annual Meeting of the Association for Computational Linguistics and the 11th International Joint Conference on Natural Language Processing (Volume 1: Long Papers), pages 4814–4828. Association for Computational Linguistics, 2021.

[3] https://www.epic.com/about/. Last access September 13, 2023.

[4] https://lhncbc.nlm.nih.gov/scrubber/. Last access September 11, 2023.

[5] Ling Luo, Shankai Yan, Po-Ting Lai, Daniel Veltri, Andrew Oler, Sandhya Xirasagar, Rajarshi Ghosh, Morgan Similuk, Peter N Robinson, and Zhiyong Lu. PhenoTagger: a hybrid method for phenotype concept recognition using human phenotype ontology. Bioinformatics, 37(13):1884–1890, 2021.

[6] Ashish Vaswani, Noam Shazeer, Niki Parmar, Jakob Uszkoreit, Llion Jones, Aidan N Gomez, Ł ukasz Kaiser, and Illia Polosukhin. Attention is all you need. In I. Guyon, U. Von Luxburg, S. Bengio, H. Wallach, R. Fergus, S. Vishwanathan, and R. Garnett, editors, Advances in Neural Information Processing Systems, volume 30. Curran Associates, Inc., 2017.

[7] L. Ouyang, J. Wu, X. Jiang, et al. Training language models to follow instructions with human feedback. In Proceedings of Advances in Neural Information Processing Systems, volume 35, pages 27730–27744, 2022.

[8] Emily Alsentzer, John Murphy, William Boag, Wei-Hung Weng, Di Jindi, Tristan Naumann, and Matthew McDermott. Publicly available clinical BERT embeddings. In Proceedings of the 2nd Clinical Natural Language Processing Workshop, pages 72–78. Association for Computational Linguistics, 2019.

[9] https://github.com/GeneDx/txt2hpo, 2019. Last access August 23, 2023.

[10] https://tedboy.github.io/nlps/generated/generated/nltk.RegexpStemmer.html. Last access October 2, 2023.

[11] Quoc Le and Tomas Mikolov. Distributed representations of sentences and documents. In Proceedings of the 31 st International Conference on Machine Learning, volume 32, 2014.

[12] Radim Ř ehůřek and Petr Sojka. Software Framework for Topic Modelling with Large Corpora. In Proceedings of the LREC 2010 Workshop on New Challenges for NLP Frameworks, pages 45–50. ELRA, 2010.

[13] https://spacy.io/universe/project/negspacy. Last access October 2, 2023.

[14] Cong Liu, Fabricio Sampaio Peres Kury, Ziran Li, Casey Ta, Kai Wang, and Chunhua Weng. Doc2Hpo: a web application for efficient and accurate HPO concept curation. Nucleic Acids Research, 47(W1):W566–W570, 2019.

[15] Aryan Arbabi, David R Adams, Sanja Fidler, and Michael Brudno. Identifying clinical terms in medical text using ontology-guided machine learning. JMIR Med Inform, 7(2):e12596, 2019.

[16] Yuhao Feng, Lei Qi, and Weidong Tian. Phenobert: A combined deep learning method for automated recognition of human phenotype ontology. IEEE/ACM Transactions on Computational Biology and Bioinformatics, 20(2):1269–1277, 2023.

[17] Scott M. Lundberg and Su-In Lee. A unified approach to interpreting model predictions. In Proceedings of the 31st International Conference on Neural Information Processing Systems, page 4768–4777. Curran Associates Inc., 2017.

[18] Bernal Jimenez Gutierrez, Nikolas McNeal, Clayton Washington, You Chen, Lang Li, Huan Sun, and Yu Su. Thinking about GPT-3 in-context learning for biomedical IE? think again. In Findings of the Association for Computational Linguistics: EMNLP 2022, pages 4497–4512, Abu Dhabi, United Arab Emirates, 2022. Association for Computational Linguistics.

[19] https://help.openai.com/en/articles/7127966-what-is-the-difference-between-the-gpt-4-models. Accessed 21 July 2023.

[20] https://www.gao.gov/products/gao-20-593. Last access September 13, 2023.

